# Single-Cell-Validated Transcriptomic Proxies for the Maas Meningioma Microenvironment Risk Continuum: An NF2-Dependent Signal Attenuated Below Detectability in Bulk RNA-seq

**DOI:** 10.64898/2026.04.27.26351840

**Authors:** Daniele Piccolo, Marco Vindigni

## Abstract

**Purpose:** Maas et al. recently showed that a microenvironment-determined risk continuum, driven by the shift from microglia-like to myeloid-derived macrophage-like tumor-associated macrophages (TAMs), independently predicts meningioma progression beyond WHO grade. Whether this gradient is recoverable from bulk RNA-seq has not been tested.

**Methods:** We computed a microglia-to-macrophage ssGSEA ratio using expanded gene sets (15 microglia, 17 macrophage) anchored to Maas core markers across 968 meningiomas from 5 GEO datasets, validated it against pseudo-bulk profiles from the Maas snRNA-seq cohort (n=25), and tested recurrence-free survival (RFS) association by Cox regression in a 101-patient subset (73 events, median follow-up 110.2 months).

**Results:** The ratio correlated with single-cell microglia proportion (overlap-controlled r=0.70, 95% CI 0.42-0.86) and discriminated WHO grades and transcriptomic clusters, confirming biological recoverability. The ratio did not predict RFS (HR 0.92, 95% CI 0.72-1.16, p=0.46). A quantitative attenuation analysis predicts the Maas IHC HR of 2.00 attenuates to HR 1.24-1.40 after proxy measurement error (r2=0.22-0.49) and NF2-wildtype dilution (30-45%), yielding only 15-40% power at 73 events. An exploratory NF2-expression proxy subgroup (uncorrected p=0.056) showed a trend in NF2-low tumors (HR 0.68, 95% CI 0.46-1.01) absent in NF2-high tumors (HR 0.98, p=0.89). The Chen 34-gene tumor-intrinsic panel also reached near-chance discrimination (C-index 0.552).

**Conclusion:** Single-cell-anchored ssGSEA recovers the Maas gradient in bulk RNA-seq but attenuates it below detectability in moderate-sized, NF2-unselected cohorts. The prognostic component is bounded by power and NF2 stratification, not an intrinsic modality failure; NF2-annotated cohorts with approximately 480 events are required for definitive testing.

## INTRODUCTION

Meningioma is the most common primary intracranial tumor, accounting for approximately 40% of all central nervous system neoplasms [1]. While the majority follow a benign clinical course, a subset recurs despite gross total resection and favorable histology. The 2021 WHO grading system, though a cornerstone of clinical decision-making, incompletely captures this biological heterogeneity [2].

Molecular approaches have progressively refined meningioma classification. The Choudhury et al. DNA methylation classification [3] established the foundational 4-group architecture from which the Maas risk continuum was derived via NMF decomposition; the Maas framework is therefore a refinement that identifies the TME as the primary biological driver of within-methylation-group risk gradients.

Transcriptomic classifiers, including the 34-gene expression panel validated by Chen et al. [4] and the four-group molecular classifier developed by Landry et al. [5], have demonstrated that molecular features add clinically actionable information beyond WHO grade. Nassiri et al. [6] proposed four molecular groups (MG1-Immunogenic, MG2-NF2 wild-type/Benign, MG3-Hypermetabolic, MG4-Proliferative) based on integrated methylation and transcriptomic data, while Vasudevan et al. [7] characterized transcriptomic subtypes within the NF2 wild-type compartment.

In a paradigm-shifting study, Maas et al. [8] introduced a microenvironment-determined risk continuum that redefines meningioma biology beyond tumor-intrinsic features. Integrating single-nucleus RNA sequencing (snRNA-seq; n=26, 44,266 nuclei), spatial transcriptomics, DNA methylation-based non-negative matrix factorization (NMF; n=4,502 methylomes), and immunohistochemistry (IHC; n=1,378), Maas et al. demonstrated that NF2-mutant meningiomas exist on a continuous spectrum defined by the composition of the tumor-associated macrophage compartment: low-risk tumors (MC ben-1) harbor predominantly microglia-like TAMs, while high-risk tumors (MC mal) are enriched in myeloid-derived proliferative macrophages [8]. In their IHC validation, low PU.1/TAM density independently predicted shorter progression-free survival after adjustment for WHO grade (HR 2.00, 95% CI 1.45-2.76, p=2.4×10^-5^, n=1,378, 321 events), and PU.1 percentage improved the C-index from 0.63 to 0.66 (p=0.0003) [8]. In a parallel methylation-based analysis, immune cell proportion predicted outcome in three independent cohorts (discovery HR 3.73, 95% CI 1.95-7.16, n=506) [8].

This finding has direct clinical implications, but whether the microglia-to-macrophage gradient is detectable in bulk RNA-seq data, the most widely available transcriptomic modality, remains untested. Standard computational deconvolution approaches estimate total macrophage burden without distinguishing microglia-like from myeloid-derived macrophage subpopulations. A prior analysis of the same cohort [9] applied three independent deconvolution methods (EPIC, MCPcounter, CIBERSORTx) and found that total macrophage scores did not predict recurrence-free survival (EPIC macrophage HR=0.90, p=0.53). This null finding is consistent with the Maas framework: if prognosis depends on the composition rather than the quantity of the myeloid compartment, total macrophage burden would be uninformative. This approach has precedent in scRNA-seq-to-bulk projection methods applied to meningioma classification [10].

We therefore hypothesized that decomposing the myeloid compartment into microglia-like and macrophage-like fractions using targeted gene sets might rescue the prognostic signal that total quantification obscures. We constructed expanded gene sets anchored to the Maas core markers, validated the resulting ssGSEA ratio against single-cell ground truth, and tested its prognostic value in the largest available public bulk RNA-seq meningioma cohort.

## METHODS

### Study Design and Data Sources

This is an in silico computational analysis using publicly available de-identified transcriptomic data. No institutional review board approval was required. This study is positioned as the single-cell-validated validation-boundary companion to the parent cohort discovery/composition analysis [9]: our previous study characterized total myeloid architecture and an estrogen-immune dissociation in this cohort; the present study provides independent single-cell-anchored validation of microglia/macrophage decomposition, derives the quantitative attenuation bounds for transcriptomic TME prognostication, and tests NF2-dependent subgroup signal. We aggregated meningioma RNA-seq data from five GEO datasets (GSE212666, GSE252291, GSE183653, GSE136661, GSE101638), yielding 968 samples across 17,585 genes after quality filtering and batch correction with limma::removeBatchEffect using GEO series as the batch variable and WHO grade as a protected biological covariate (PERMANOVA-verified residual batch variance R^2^<0.05) [9]. The expression matrix is in log_2_(TPM+1) scale.

For single-cell validation, the Maas et al. snRNA-seq Seurat object (44,266 nuclei, 26 samples) was obtained from their Zenodo deposit (doi:10.5281/zenodo.17296825) [8].

NF2 mutation status was not annotated in the available bulk RNA-seq metadata and could not be reliably inferred from expression data alone. The Maas risk continuum is defined within NF2-mutant meningiomas [8]; our unselected cohort therefore includes an estimated 30-40% NF2-wild-type tumors in which the microglia-to-macrophage gradient may not apply.

### Microglia and Macrophage Gene Set Construction

Two gene sets were constructed to capture the microglia-like versus myeloid-derived macrophage gradient described by Maas et al. [8]. The microglia gene set (15 genes) comprised the five Maas core markers (TMEM119, P2RY12, P2RY13, GPR34, SLC2A5) plus ten established microglial identity genes (CX3CR1, HEXB, SALL1, CSF1R, TREM2, OLFML3, SELPLG, SPI1, SIGLEC8, AIF1) [11, 12]. The macrophage gene set (17 genes) comprised the five Maas peripheral-macrophage markers (C3, F10, EMILIN2, F5, GDA) plus HP and eleven activated/tumor-supportive genes enriched in high-risk MC classes (SPP1, CD163, MARCO, FOLR2, MSR1, CD68, ITGAM, CCL2, TGFB1, IL10, CD83) [8, 13, 14]. MKI67, TOP2A (tumor-proliferation confounders [15]) and SELL (lymphocyte marker) were excluded; a sensitivity analysis including these was performed. Full gene-level justification, including TREM2 bidirectional roles, SPI1 pan-myeloid expression, and SIGLEC8 substitution for the absent SIGLECH, is in Supplementary Table S1.

### Single-Sample Gene Set Enrichment Analysis

Per-sample enrichment scores were computed for each gene set using single-sample gene set enrichment analysis (ssGSEA) implemented in the GSVA R package (v2.4.4) [16] with default parameters and normalization enabled. The microglia-to-macrophage ratio was defined as:

*Ratio = ssGSEAmicroglia - ssGSEAmacrophage*

Higher values indicate microglia dominance (lower risk per the Maas continuum); lower values indicate macrophage dominance (higher risk). All ratio scores were z-scored across the full cohort (mean=0, SD=1) prior to downstream analysis.

As a complementary scoring method, mean z-scores of gene expression within each gene set were computed, and the ratio was calculated analogously. Both methods were carried forward to assess scoring method sensitivity.

### Pseudo-Bulk Validation Against Single-Cell Ground Truth

To validate that the ssGSEA ratio captures the Maas microglia-to-macrophage gradient, we performed pseudo-bulk validation using the Maas snRNA-seq cohort [8]. From the Seurat object (44,266 cells, 26 samples), 15,779 cells annotated as macrophages (TAMs) were extracted. Each TAM was classified as microglia-like or macrophage-like based on higher normalized expression of the microglia versus macrophage core gene sets, respectively.

Pseudo-bulk expression profiles were generated by summing raw counts across all cell types per sample (matching the mixed-cell-type composition of bulk RNA-seq) and converting to log_2_(CPM+1). The ssGSEA ratio was computed on these pseudo-bulk profiles. Samples with fewer than 10 TAMs were excluded, yielding 25 evaluable samples.

The primary validation metric was the Spearman rank correlation between the pseudo-bulk ssGSEA ratio and the single-cell microglia proportion. A secondary metric was the correlation with Maas MC risk class (coded numerically: ben-1=1, ben-2=2, int-A=3, int-B=4, mal=5). The pre-specified pass criterion was |r| >= 0.40 with p < 0.05.

### Survival Analysis

The primary survival cohort comprised 101 patients from the previously described survival subset [9] with complete data for recurrence-free survival (time and event), WHO grade, and age (73 recurrence events; median follow-up 110.2 months by reverse Kaplan-Meier). This cohort was composed of Baylor/GSE136661 (n=17), UCSF 2022 (n=15), and UW/FHCC (n=69). WHO grade distribution was grade I (n=34), grade II (n=52), grade III (n=15) (Supplementary Table S2).

WHO grade was entered as a single ordinal covariate (numeric: 1, 2, 3), assuming monotonic association with hazard. Factor-coded sensitivity analysis yielded equivalent results (data not shown).

The events-per-variable (EPV) ratio for the primary two-term model (ratio + WHO grade) was 36.5, exceeding the conventional threshold of 10. With 73 events, the 95% confidence interval width (0.72-1.16) excludes effects larger than HR 1.16 per SD, providing adequate power to detect effects of the magnitude reported by Maas et al. (HR 2.00 for IHC-based PU.1 quantification).

The primary analysis was a univariable Cox proportional hazards model with the z-scored ssGSEA ratio as a continuous predictor (HR per 1 SD). A multivariable model additionally adjusted for WHO grade (ordinal, 1/2/3). The proportional hazards assumption was verified using Schoenfeld residual tests. Kaplan-Meier curves were generated after stratifying patients by pre-specified ratio tertiles (low, intermediate, high), and differences were assessed by log-rank test. Tertile cutpoints were fixed from the full 968-sample cohort to prevent data-adaptive selection within the survival subset.

C-index comparison quantified the discriminative improvement of adding the ratio to WHO grade alone. Three models were compared: (A) WHO grade + total macrophage score (from prior deconvolution [9]), (B) WHO grade + microglia/macrophage ratio, and (C) WHO grade + ratio + total macrophage score. The Wald test p-value for each TME variable within its respective multivariable model assessed the incremental value over WHO grade. Permutation testing (10,000 permutations of the ratio label) estimated the empirical significance of the observed Cox z-statistic.

Sensitivity analyses tested alternative scoring methods: mean z-score ratio, and ssGSEA ratio with proliferation markers included. Both univariable and multivariable (adjusted for WHO grade) results were computed for each method.

Extended sensitivity analyses (Supplementary Table S6) addressed potential confounders: (a) age adjustment (WHO + age + ratio); (b) institution-stratified Cox with strata(dataset) allowing institution-specific baseline hazards; (c) NF2 expression proxy analysis, stratifying the survival cohort by median NF2 mRNA expression (NF2-low enriched for NF2-mutant tumors) and repeating the primary Cox model in each subset; (d) composition-corrected ratio, residualizing the ssGSEA ratio against total macrophage score (EPIC) to isolate the composition-specific component; (e) microglia and macrophage ssGSEA scores tested individually as survival predictors; (f) the Chen et al. 34-gene expression panel [4] computed as a single ssGSEA score and tested as a benchmark for RNA-seq prognostication in this cohort; (g) formal paired C-index comparison using the compareC package [17].

### Framework Concordance

To contextualize the microglia/macrophage ratio within existing molecular classification frameworks, its distribution was assessed across two reference schemes. Transcriptomic cluster assignments for a 225-sample subset were inherited from the parent study [9]. These seven clusters (A-G) were defined by Thirimanne et al. [18] through unsupervised consensus clustering of the full expression matrix, capturing immune-enriched (Cluster B), immune-desert (Cluster C), and proliferative (Clusters E-G) subtypes.

Nassiri molecular group (MG) proxies were constructed by computing ssGSEA scores for MSigDB Hallmark gene sets [19] corresponding to each Nassiri MG [6]: MG1-Immunogenic (inflammatory response, allograft rejection, interferon-gamma response, IL6-JAK-STAT3, complement), MG2-NF2wt/Benign (angiogenesis, epithelial-mesenchymal transition, coagulation), MG3-Hypermetabolic (fatty acid metabolism, oxidative phosphorylation, adipogenesis, bile acid metabolism), and MG4-Proliferative (E2F targets, G2M checkpoint, MYC targets V1, mitotic spindle). Each sample was assigned to the MG with the highest mean constituent pathway score.

A pre-specified three-way framework concordance analysis (Maas × Vasudevan × Landry/Nassiri) could not be executed as planned: the Nassiri/Landry group-specific signatures were not available to us at the time of this analysis, and a Hallmark-pathway-based proxy failed distribution sanity checks (69% of samples assigned to MG3-Hypermetabolic) and is reported in Supplementary Table S5 as a methodological negative. Vasudevan subtype assignments additionally require NF2 status annotation not present in our cohort. The Kruskal-Wallis test assessed whether the microglia/macrophage ratio differed across groups in each framework. Effect sizes were quantified using eta-squared (η^2^).

### Leave-One-Dataset-Out Stability Analysis

To test the institutional stability of the microglia/macrophage ratio, a leave-one-dataset-out (LODO) analysis was performed across the five constituent GEO datasets. For each iteration, one dataset was held out. The ssGSEA ratio was recomputed on the remaining samples and z-scored. Spearman rank correlation between the original and LODO-recomputed ratio was the stability metric. A dataset was flagged as an outlier if its exclusion shifted the mean ratio by more than 0.5 SD.

Because the underlying batch-corrected matrix was computed once using all samples, LODO within this matrix has residual information leakage from shared batch correction parameters, constraining rank-order changes and inflating stability estimates. The pseudo-bulk validation on the independent Maas snRNA-seq cohort serves as the leakage-free external validation.

### Statistical Analysis

All analyses were performed in R (v4.5.0). ssGSEA scores were computed with the GSVA package (v2.4.4) [16]. Survival analysis used the survival [20] and survminer packages. C-index comparisons used Harrell’s concordance statistic. All statistical tests were two-sided. Significance was set at alpha=0.05 for pre-specified primary analyses. Bonferroni correction was applied for WHO grade subgroup analyses (alpha=0.025 per stratum). Supplementary Table S6 sensitivity analyses are reported as exploratory with uncorrected p-values; inferential claims rest on the pre-specified primary analyses only. The proportional hazards assumption was assessed via Schoenfeld residual tests and was satisfied for all models (global *p*>0.05). Analysis code will be deposited on GitHub upon publication.

## RESULTS

### The Ratio Does Not Predict Recurrence-Free Survival

The primary outcome of this study was negative. In the 101-patient survival cohort (73 events), the microglia-to-macrophage ssGSEA ratio did not predict recurrence-free survival (Table 1).

**Table 1.** Cox Proportional Hazards Models for Recurrence-Free Survival.

| Model | Variable | HR | 95% CI | p |
| --- | --- | --- | --- | --- |
| Univariable | Ratio (per SD) | 0.92 | 0.72-1.16 | 0.46 |
| Multivariable | Ratio (per SD) | 0.98 | 0.77-1.24 | 0.85 |
|  | WHO grade (ordinal) | 1.73 | 1.19-2.51 | 0.004 |
| WHO grade only | WHO grade (ordinal) | 1.74 | 1.21-2.51 | 0.003 |

In univariable Cox regression, the ratio showed a non-significant trend (HR 0.92 per SD, 95% CI 0.72-1.16, *p*=0.46). The ratio alone had near-chance discriminative ability (C-index 0.551, 95% CI 0.461-0.630; Table 2). Kaplan-Meier analysis by ratio tertiles showed no survival separation (log-rank *p*=0.79; Fig. 1A). In the multivariable model adjusting for WHO grade, the ratio contributed no independent prognostic information (HR 0.98, 95% CI 0.77-1.24, *p*=0.85), while WHO grade remained significant (HR 1.73, 95% CI 1.19-2.51, *p*=0.004).

**Figure 1.**
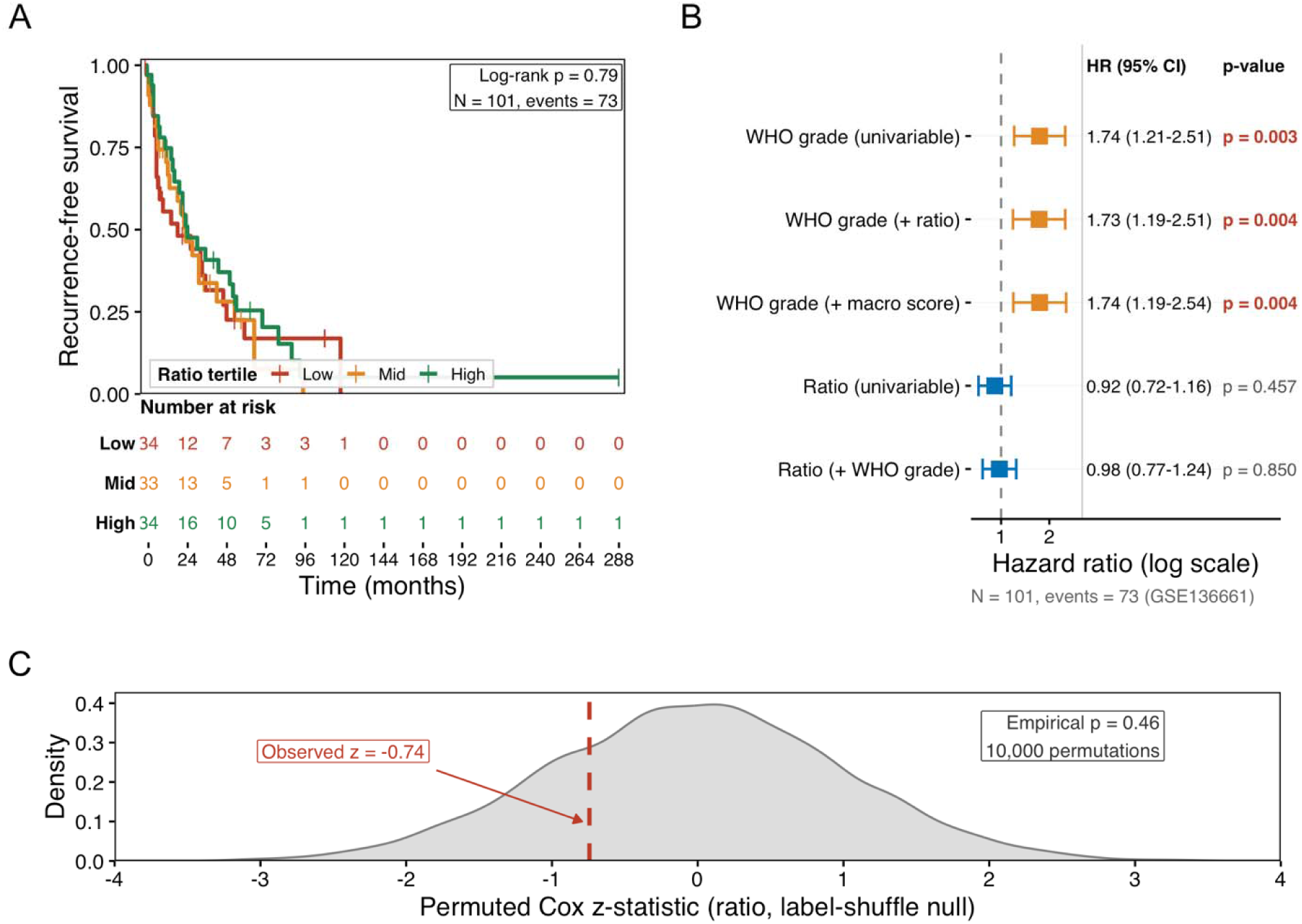
(A) Kaplan-Meier curves for recurrence-free survival stratified by ratio tertiles (low, intermediate, high). Log-rank p=0.79. **(B)** Forest plot of Cox regression hazard ratios for all models tested: ratio univariable, ratio + WHO multivariable, WHO-only reference, and prior deconvolution total macrophage + WHO reference [9]. **(C)** Permutation null distribution (10,000 permutations) of the Cox z-statistic with the observed value indicated. Empirical *p*=0.46.

**Table 2.** C-Index Comparison Across Prognostic Models.

| Model | C-index | 95% CI | AIC | Wald p for TME term |
| --- | --- | --- | --- | --- |
| Ratio alone (univariable) | 0.551 | 0.461-0.630 | — | 0.46 |
| WHO grade only | 0.594 | 0.513-0.663 | 532.5 | — |
| WHO + ratio | 0.616 | 0.523-0.688 | 534.5 | 0.85 |
| WHO + macro score | 0.576 | 0.524-0.692 | 534.5 | 0.98 |
| WHO + ratio + macro | 0.614 | 0.534-0.694 | 536.5 | — |

N=101, 73 events. C-index = Harrell's concordance statistic. Wald p for TME term = p-value for the TME variable within each multivariable model, testing incremental value over WHO grade. Macro score = total macrophage score from prior deconvolution analysis. LRT for ratio adding to WHO grade alone: $p=0.85$ . Formal paired C-index comparison (compareC): WHO vs. WHO+ratio delta-C = +0.022, $z=1.27$ , $p=0.20$ ; WHO vs. WHO+macro delta-C = -0.018, $z=-0.99$ , $p=0.32$ .

In a pre-specified head-to-head comparison [Methods §2.6], adding the ratio to WHO grade alone improved discrimination from C-index 0.594 to 0.616 (paired compareC ΔC = +0.022, z = 1.27, *p* = 0.20), while substituting the total macrophage score from prior deconvolution [9] reduced discrimination (C-index 0.576, ΔC = −0.018, *p* = 0.32; Table 2).

A forest plot of all Cox models tested is shown in Fig. 1B. Permutation testing with 10,000 label permutations yielded an empirical p-value of 0.46 (95% CI 0.45-0.47; Fig. 1C), confirming the null finding. All models satisfied the proportional hazards assumption (Schoenfeld global test: univariable *p*=0.28; multivariable *p*=0.61).

The null finding persisted across all scoring methods in both univariable and multivariable analyses (all *p*>0.33 univariable, all *p*>0.60 multivariable; Table 3).

**Table 3.** Sensitivity Analysis: Survival Results Across Scoring Methods.

| Method | HR (univar) | 95% CI | p (univar) | HR (MV+WHO) | 95% CI | p (MV) |
| --- | --- | --- | --- | --- | --- | --- |
| ssGSEA ratio (primary) | 0.92 | 0.72-1.16 | 0.46 | 0.98 | 0.77-1.24 | 0.85 |
| Mean z-score ratio | 0.95 | 0.76-1.19 | 0.63 | 1.06 | 0.84-1.34 | 0.62 |
| ssGSEA + prolif. markers | 0.89 | 0.70-1.13 | 0.34 | 0.98 | 0.76-1.26 | 0.85 |
| Mean z + prolif. markers | 0.91 | 0.72-1.14 | 0.40 | 1.03 | 0.81-1.31 | 0.81 |

N=101, 73 events. HR per 1 SD increase in ratio score. MV = multivariable model adjusted for WHO grade (ordinal).

Extended sensitivity analyses adjusting for age, stratifying by institution, testing composition-corrected ratios, and testing individual microglia and macrophage component scores all yielded null results (Supplementary Table S6). As a benchmark, the Chen et al. 34-gene prognostic panel [4] (32/34 genes available) — validated at AUC 0.81 in 1,856 patients — also achieved near-chance discrimination in this cohort (C-index 0.552, 95% CI 0.46-0.63; HR 0.92, *p*=0.49; Supplementary Table S6).

### The Ratio Captures the Maas Gradient in Single-Cell Data

To validate the ssGSEA ratio as a proxy for the Maas microglia-to-macrophage gradient, we analyzed pseudo-bulk profiles from the Maas snRNA-seq cohort. Among 15,779 TAMs across 25 evaluable samples, 6,234 (39.5%) were classified as microglia-like and 9,545 (60.5%) as macrophage-like based on core marker expression.

The single-cell microglia proportion decreased monotonically from low-risk to high-risk MC classes: ben-1, 47.7%; ben-2, 36.5%; int-A, 32.8%; int-B, 35.6%; mal, 14.4% — and from WHO grade 1 (41.9%) through grade 2 (25.4%) to grade 3 (24.2%) (Supplementary Table S3). The pseudo-bulk ssGSEA ratio correlated strongly with the single-cell microglia proportion when cells were classified using the expanded gene sets (Spearman r=0.77, 95% CI 0.54-0.89, *p*=6.6×10^-6^; Fig. 2A), exceeding the pre-specified validation threshold of |r|>=0.40.

**Figure 2.**
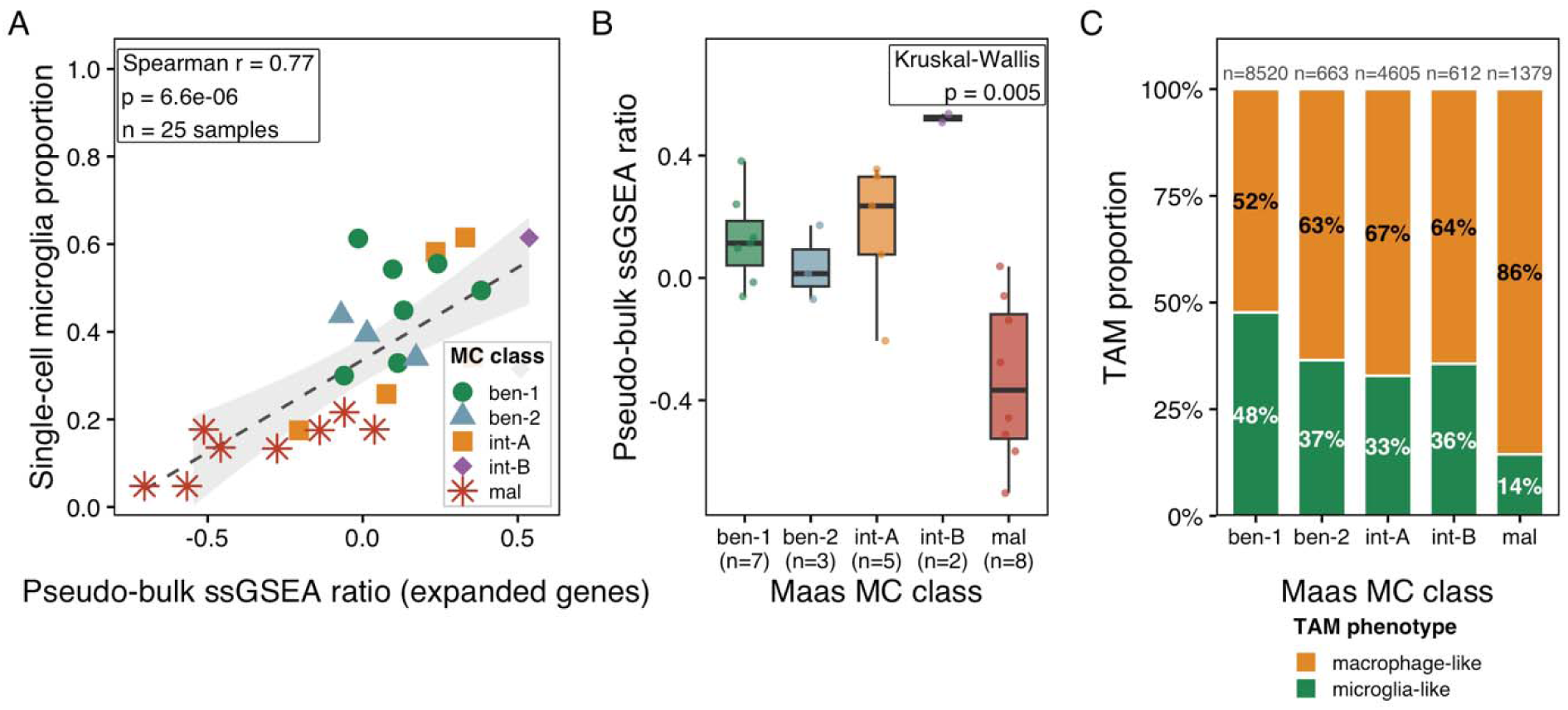
Pseudo-bulk validation of the microglia/macrophage ssGSEA ratio. (A) Scatter plot of the pseudo-bulk ssGSEA ratio versus single-cell microglia proportion across 25 Maas snRNA-seq samples, colored by MC class. Spearman r=0.77, *p*=6.6×10^-6^. (B) Box plot of the ssGSEA ratio by Maas MC class. The ratio decreases from ben-1 to mal, consistent with the microenvironment risk continuum. Kruskal-Wallis *p*=0.005. int-B (n=2) is displayed but unreliable. (C) Stacked bar plot showing the proportion of microglia-like versus macrophage-like TAMs by MC class and WHO grade.

Because the expanded gene sets overlap between the cell-level classification and the ssGSEA scoring, we performed a circularity analysis using the Maas core markers only (5 microglia + 6 macrophage genes) for cell classification: the core-only classification yielded a non-significant correlation (r=-0.24, *p*=0.25), while a strict classification removing dual-expression genes (TREM2, CSF1R, SPI1, SIGLEC8) yielded r=0.70 (*p*=0.0001). The conservative estimate from the strict classification confirms that the ssGSEA ratio captures real biological signal, though the r=0.77 estimate is inflated by gene set overlap.

The ratio also correlated with the Maas MC risk class, though more modestly (r=-0.47, 95% CI −0.73 to - 0.10, *p*=0.017), reflecting the expected information loss from continuous biology to ordinal classification. The pseudo-bulk ssGSEA ratio thus captures approximately 22% of variance in the MC risk class (r^2^=0.22), quantifying the measurement error inherent to the bulk RNA-seq proxy. Mean ssGSEA ratio by MC class was: ben-1, +0.13; ben-2, +0.04; int-A, +0.16; int-B, +0.52; mal, −0.33 (Kruskal-Wallis *p*=0.005; Fig. 2B). The int-B class showed an unexpectedly high ratio (n=2 samples only, unreliable); excluding int-B, the gradient from ben-1 to mal was monotonically decreasing.

### The Ratio Associates with Tumor Biology in the Full Cohort

In the full 968-sample bulk RNA-seq cohort, the z-scored ratio exhibited adequate inter-sample variance (IQR=1.35). The ratio correlated significantly with WHO grade in the 160-sample subset with grade annotation (Spearman r=-0.19, *p*=0.018; Fig. 3A), with mean ratio decreasing from grade I (+0.04, n=121) through grade II (−0.28, n=32) to grade III (−1.02, n=7). Higher-grade tumors showed greater macrophage dominance, consistent with the Maas framework.

**Figure 3.**
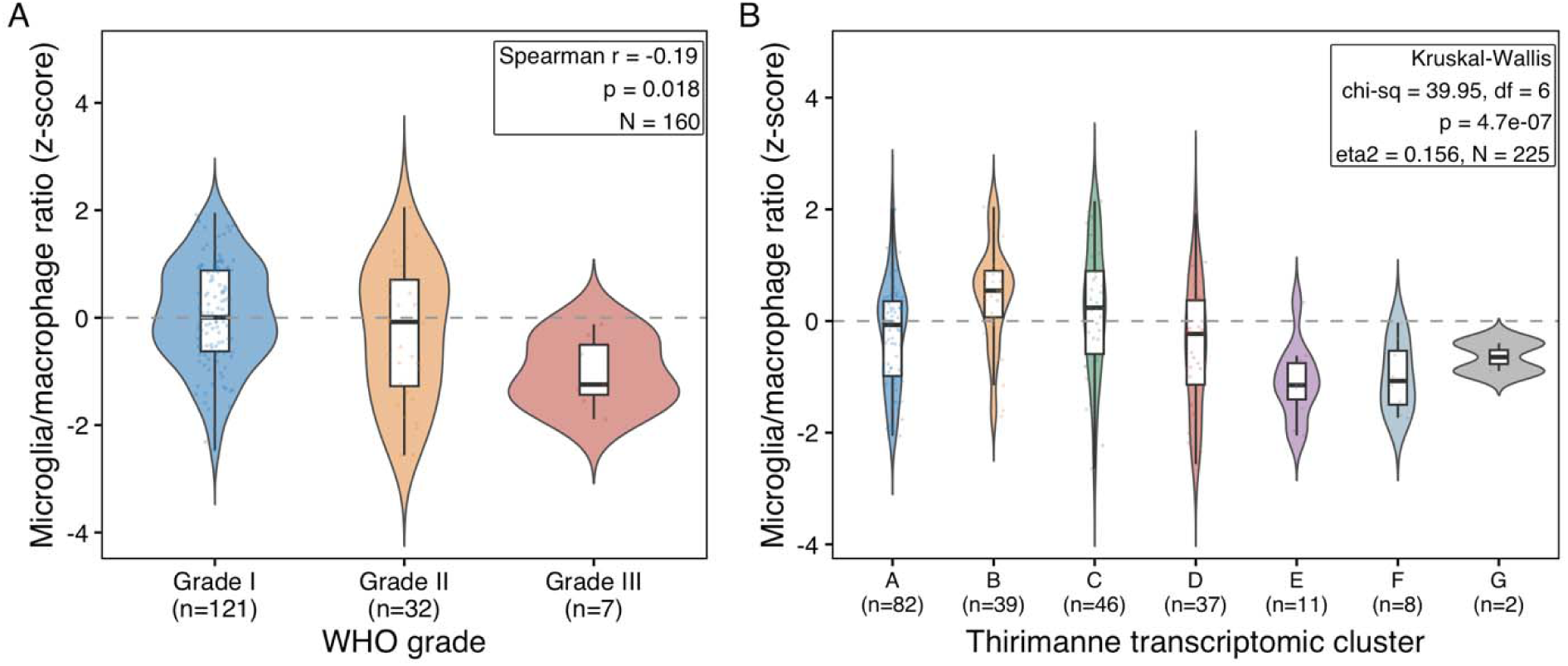
Microglia/macrophage ratio distribution in the full bulk RNA-seq cohort (N=968). (A) Box plot of the z-scored ratio by WHO grade (N=160 with grade annotation). Spearman r=-0.19, *p*=0.018. (B) Box plot of the z-scored ratio by transcriptomic cluster (N=225). Kruskal-Wallis *p*=4.7×10^-7^, η^2^=0.156.

Among the 225 samples with transcriptomic cluster assignments, the ratio differed significantly across seven clusters (Kruskal-Wallis chi^2^=39.95, df=6, *p*=4.7×10^-7^; η^2^=0.156; Fig. 3B). The ratio showed negligible association with dataset of origin (Kruskal-Wallis η^2^ < 0.01 across the 5 GEO accessions), confirming that the biological signal is independent of batch.

The ratio was moderately correlated with total macrophage scores from the prior deconvolution analysis [9] (Spearman r=0.24 with EPIC macrophages, r=0.36 with MCPcounter monocytic lineage), indicating partially overlapping but distinct biological information.

### Stability Across Datasets and Scoring Methods

LODO analysis showed no evidence of dataset-specific artifacts (mean ratio shift <0.11 SD across all iterations; Table 4; Fig. 4A), though the shared batch correction constrains rank-order changes and the r=1.0 correlations are expected given the deterministic nature of ssGSEA on a jointly preprocessed matrix. Scoring method sensitivity analysis showed high inter-method agreement (Fig. 4B): ssGSEA vs. mean z-score, r=0.94; ssGSEA vs. ssGSEA with proliferation markers, r=0.97; mean z-score vs. mean z-score with proliferation markers, r=0.98 (Table 4).

**Figure 4.**
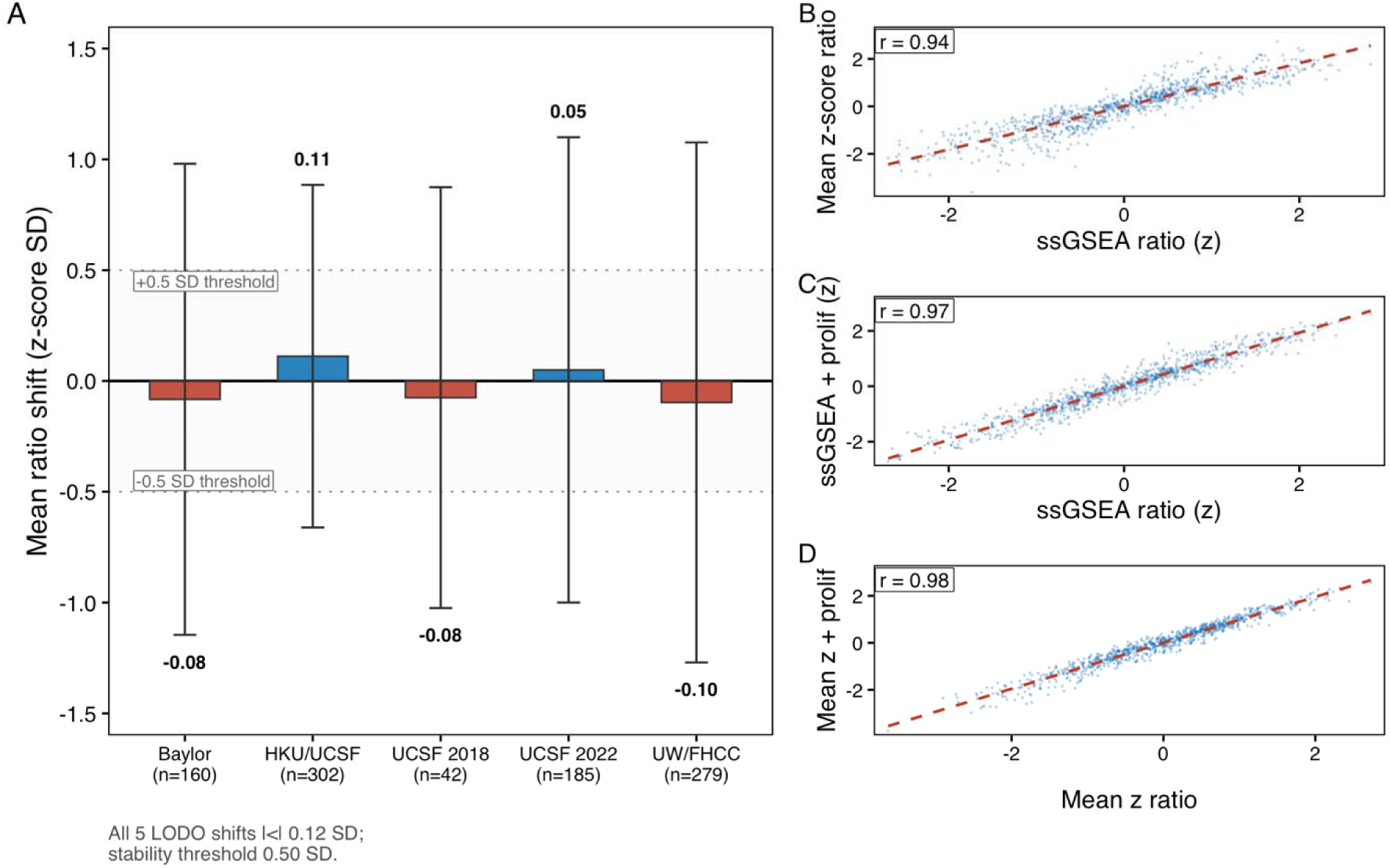
LODO stability and scoring method sensitivity. (A) Bar plot of mean ratio in held-out datasets across five LODO iterations. All mean shifts <0.11 SD. (B) Scatter matrix of ratio scores computed by four alternative scoring methods (ssGSEA, mean z-score, ssGSEA with proliferation markers, mean z with proliferation markers). All pairwise r>=0.92.

**Table 4.**
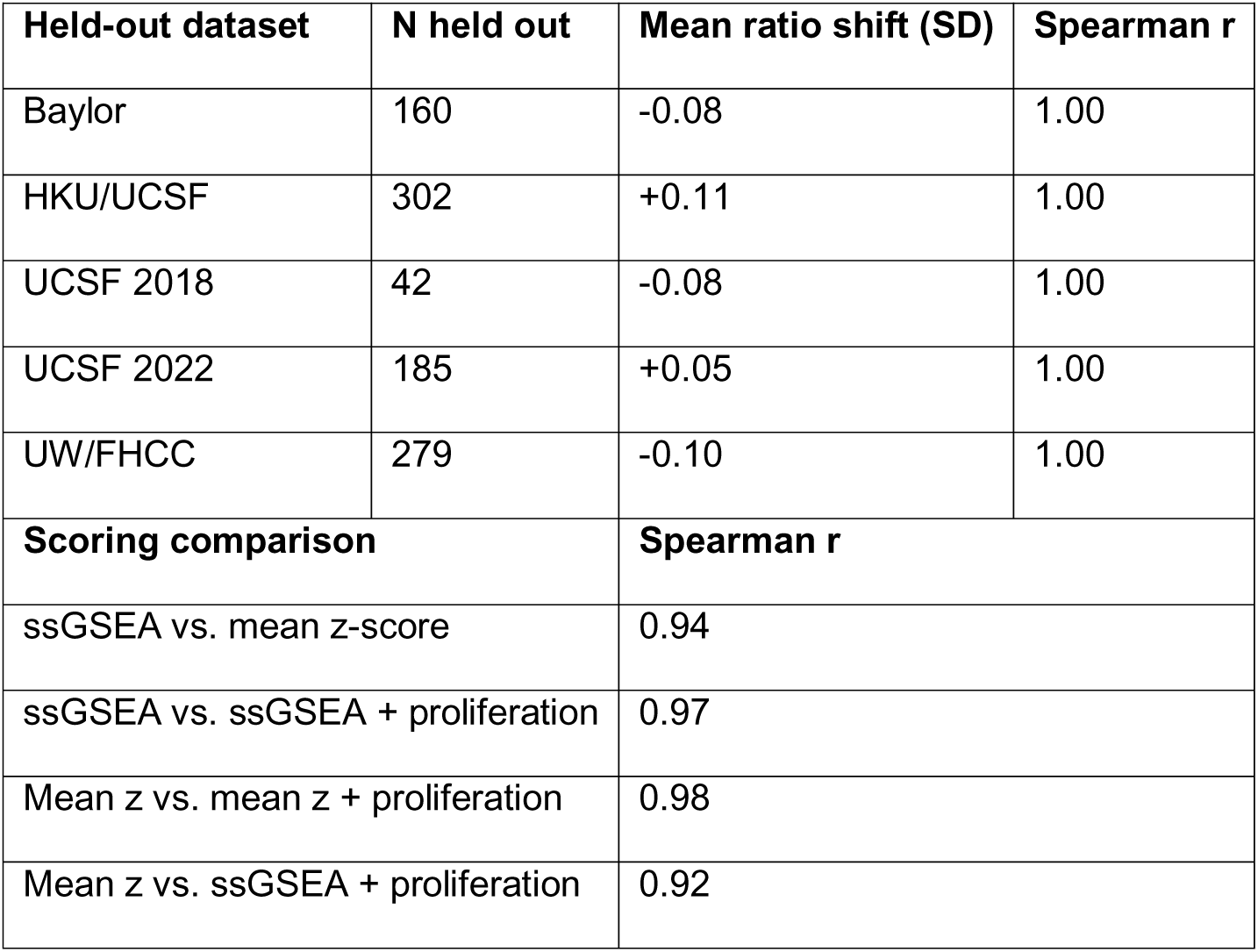
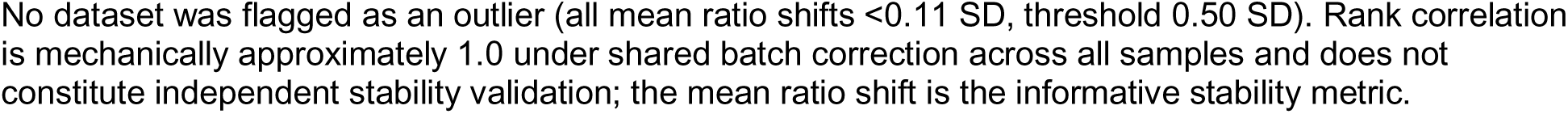
LODO Stability and Scoring Method Sensitivity.

| Held-out dataset | N held out | Mean ratio shift (SD) | Spearman r |
| --- | --- | --- | --- |
| Baylor | 160 | -0.08 | 1.00 |
| HKU/UCSF | 302 | +0.11 | 1.00 |
| UCSF 2018 | 42 | -0.08 | 1.00 |
| UCSF 2022 | 185 | +0.05 | 1.00 |
| UW/FHCC | 279 | -0.10 | 1.00 |
| Scoring comparison |  | Spearman r |  |
| ssGSEA vs. mean z-score |  | 0.94 |  |
| ssGSEA vs. ssGSEA + proliferation |  | 0.97 |  |
| Mean z vs. mean z + proliferation |  | 0.98 |  |
| Mean z vs. ssGSEA + proliferation |  | 0.92 |  |

No dataset was flagged as an outlier (all mean ratio shifts <0.11 SD, threshold 0.50 SD). Rank correlation is mechanically approximately 1.0 under shared batch correction across all samples and does not constitute independent stability validation; the mean ratio shift is the informative stability metric.

### WHO Grade Subgroup Analysis

The ratio decreased monotonically across WHO grades (grade I: +0.04, n=121; II: −0.28, n=32; III: −1.02, n=7), with the largest shift between grades 1 and 2 (microglia proportion: 41.9% to 25.4%). Within WHO grade 2 — the largest survival-cohort subgroup (n=52, 45 events) and the stratum with the strongest Maas PU.1 effect (HR 2.91) [8] — the ratio showed no survival discrimination (log-rank *p*=0.63).

## DISCUSSION

### Signal Attenuation Is Quantitatively Predictable

The null RFS association for the microglia/macrophage ratio is not a surprise but a prediction. Applying classical errors-in-variables attenuation to the Maas IHC reference (HR = 2.00 for PU.1/TAM density, 95% CI 1.45-2.76 at n=1,378 events=321 [8]), the expected observable HR in bulk RNA-seq depends on two compounding factors: (i) the reliability of the ssGSEA ratio as a proxy for the Maas microenvironment risk class, and (ii) the proportion of NF2-wildtype tumors diluting an NF2-specific effect. Under plausible reliability estimates (r^2^ = 0.22 from the ordinal MC-class correlation; r^2^ = 0.49 from the strict pseudo-bulk microglia-proportion correlation) and a 30-45% NF2-wildtype grade-weighted dilution [3, 8], the expected observable HR ranges from approximately 1.24 to 1.40, corresponding to 15-40% power at 73 events and alpha=0.05 (Supplementary Methods, Attenuation Analysis Derivation). The paired compareC comparison had approximately 40% power to detect a ΔC of +0.03 and 80% power for +0.06 at 73 events and alpha=0.05 [17]; the observed +0.022 is therefore consistent with either no improvement or a small improvement below the detection floor.

Consistent with this quantitative prediction, an exploratory NF2 expression proxy analysis (uncorrected *p*-values) revealed a differential effect: in the NF2-low subset (enriched for NF2-mutant tumors; n=51, 36 events; EPV=18 per covariate), the ratio showed a trend toward prognostic value (HR 0.68, 95% CI 0.46-1.01, *p*=0.056), while in the NF2-high subset (n=50, 37 events) the ratio was null (HR 0.98, *p*=0.89). This directional differential is the falsification test that the attenuation model predicts: the signal is expected to survive preferentially in NF2-mutant tumors, and it does. The exploratory character of this subgroup analysis and the imperfect NF2 mRNA proxy preclude inferential claims, but the finding is hypothesis-generating and directly motivates NF2-stratified validation.

### Biological Validity of the Recovered Signal

The pseudo-bulk validation provides strong evidence that the Maas microglia-to-macrophage gradient is recoverable from bulk RNA-seq. The correlation between the ssGSEA ratio and the true single-cell microglia proportion reached r=0.77 with the expanded gene sets and r=0.70 (95% CI 0.42-0.86 by Fisher z-transform, n=25, *p*=0.0001) with a circularity-controlled strict classification, substantially exceeding what would be expected from a non-specific inflammatory signal. In the full bulk RNA-seq cohort, the ratio discriminated between WHO grades (Spearman rho=-0.19, *p*=0.018) and between transcriptomic clusters with substantial effect size (η^2^=0.156). The variance across dataset of origin was negligible (Kruskal-Wallis η^2^ < 0.01), confirming biological rather than technical origin of the signal. The pseudo-bulk ratio captured approximately 22-49% of variance in the Maas MC risk class (r^2^ range across classification specifications), quantifying the measurement error inherent to the bulk RNA-seq proxy. The signal is recovered but is small relative to the noise introduced by the bulk-averaging step.

### Decomposition vs Quantity: What the Ratio Measures

The Maas PU.1 IHC metric and our ssGSEA ratio measure different biological quantities. PU.1% reflects total TAM density as a fraction of all cells, essentially a spatial abundance measure, whereas the ssGSEA ratio measures the relative balance between microglia-like and macrophage-like transcriptional programs within the myeloid compartment. The prognostic value of PU.1% may therefore derive primarily from total TAM density rather than myeloid composition per se. We tested the microglia ssGSEA score alone (a closer analog to PU.1%) and found it was also non-prognostic (Supplementary Table S6), indicating that the null finding is not attributable to metric construction alone.

The magnitude of the underlying modality limitation is quantified directly by the companion analysis [9], which computed ssGSEA scores for the Maas microglia-like marker set and the Maas peripheral-macrophage marker set on the same N=968 cohort and correlated them with three consensus deconvolution estimates. The consensus macrophage score correlated strongly with the Maas microglia-like signature (Spearman rho = 0.58-0.73 across EPIC, MCPcounter, and CIBERSORTx) but only weakly with the Maas peripheral-macrophage signature (rho = 0.14-0.35). This asymmetry demonstrates that standard bulk RNA-seq deconvolution tools, whose reference matrices derive from peripheral blood, predominantly capture the brain-resident microglia compartment and only weakly register the peripheral-BMDM infiltration whose expansion drives the Maas NF2-adjusted risk continuum. The null RFS finding is therefore not contradictory to Maas 2026 but is a direct, quantitatively predictable consequence of this reference-matrix mismatch: the population whose quantitative shift is prognostic in IHC is precisely the one that bulk deconvolution under-detects. The present finding complements the coarse-quantification null reported in the companion cohort analysis [9]. Decomposition of the myeloid compartment into microglia-like and macrophage-like fractions preserves the direction of attenuation rather than rescuing the signal, consistent with a structural rather than analytical limitation of bulk transcriptomic averaging. The ssGSEA ratio showed a modest positive correlation with total macrophage scores (Spearman rho=0.24 with EPIC macrophages); a composition-corrected analysis residualizing the ratio against total macrophage burden yielded an equivalent null (Supplementary Table S6), confirming the null reflects the composition-specific component and not a quantity confound.

### Why IHC Works Where RNA-seq Does Not

The measurement modalities differ fundamentally. Maas et al. validated the prognostic association using IHC protein quantification (PU.1) on tissue microarrays and methylation-based deconvolution (Edec), both of which preserve spatial and cell-density information lost in bulk RNA-seq averaging. Our findings align with those of Lotsch et al. [13], who demonstrated that CD68-positive TAM density by immunohistochemistry independently predicted poor survival in 680 meningiomas (HR 2.11), and with the Maas PU.1 IHC result (HR 2.00) [8]. Immunohistochemistry captures spatial abundance of specific cell populations at protein level, while bulk RNA-seq deconvolution estimates fractional composition from a mixed transcriptome. Prognostically relevant macrophage populations may be spatially concentrated at the brain-tumor interface [21], a region that may be underrepresented in bulk RNA-seq from resected specimens. Guo et al. [15] further demonstrated that Ki-67 in meningioma is contributed by both tumor and immune cells, suggesting that standard Ki-67-based prognostication may itself partly capture the TME gradient characterized by Maas et al. A biological caveat qualifies the “microglia-like” language. TMEM119 and P2RY12, the canonical microglial identity markers in our set, were originally characterized in parenchymal brain microglia. Meningioma is an extra-axial tumor arising from arachnoid cap cells in the subarachnoid space, and the TMEM119+/P2RY12+ cells detected here may include pia-adjacent border-associated macrophages (BAMs) that share substantial transcriptional identity with parenchymal microglia but arise from a distinct anatomical niche. The Aggarwal et al. spatial analysis of the meningioma brain-tumor interface [21] supports this interpretation: microglia-like transcriptional programs are enriched at the tumor-parenchyma boundary. For the purposes of this analysis, “microglia-like” denotes transcriptional similarity to the Maas microglia-annotated TAM cluster rather than confirmed parenchymal microglia identity, consistent with the conceptual framing of Maas et al. [8]. Disambiguating microglia proper from BAMs would require spatial transcriptomics or BAM-specific markers such as LYVE1, F13A1, or MRC1, which are not resolvable in our bulk RNA-seq design.

### Limitations

Several limitations temper these conclusions. First, NF2 mutation status was unavailable; the Maas continuum is specifically defined for NF2-mutant meningiomas, and an unselected cohort dilutes NF2-specific signal. NF2 mRNA is an imperfect proxy — missense mutations and chr22q heterozygous loss may not reduce transcript abundance, and Merlin inactivation can occur post-translationally via S13 dephosphorylation [22]; a chr22q-loss expression-burden proxy would be more faithful but is deferred to NF2-annotated cohorts. Second, the survival cohort is small (n=101, 73 events) and dominated by UW/FHCC (68.3%); an institution-stratified Cox model yielded concordant null results (Supplementary Table S6). Third, extent of resection, the strongest modifiable predictor of meningioma recurrence [23, 24], was not available as a covariate; if TME composition correlates with resectability, EOR confounding could contribute to the null. Fourth, the gene sets are expanded from Maas core markers using literature-derived genes, not the exact Maas signatures; our approach tests the transcriptomically resolvable component, not the full multi-modal architecture. Fifth, the pseudo-bulk validation used only 25 samples, and SIGLEC8 in our microglia set substitutes for the intended but absent SIGLECH. Sixth, the pre-specified three-way concordance (Maas × Vasudevan × Landry/Nassiri) could not be executed as planned: the Nassiri/Landry signatures were not available to us at the time of this analysis, and the Hallmark-proxy distributional failure is reported as a methodological negative (Supplementary Table S5). Seventh, LODO stability analysis carries residual information leakage from shared batch correction; the pseudo-bulk analysis serves as the leakage-free external validation.

## Conclusions

This study establishes three quantitative boundaries for transcriptomic interrogation of the meningioma microenvironment. First, at the single-cell level, the ssGSEA ratio recovers the Maas microglia-to-macrophage gradient with a conservative correlation of r=0.70 against ground truth, a measurable signal. Second, at the bulk cohort level, the transcriptomic proxy captures approximately 22-49% of MC risk class variance, attenuating the reference IHC effect size from HR 2.00 to approximately 1.24-1.40 after NF2-wildtype dilution. Third, at the study-design level, detection of this attenuated effect requires approximately 480 events at 80% power, an order of magnitude beyond the 73 events available in the current public bulk RNA-seq meningioma literature. Three forward-looking design choices follow: (i) NF2-stratified bulk RNA-seq cohorts of approximately 500 events, the only approach that could rescue the signal at the transcriptomic layer; (ii) protein-level assays such as PU.1 IHC, which preserve the spatial and density information lost by bulk averaging and are the Maas reference modality; and (iii) spatial transcriptomic approaches (CytoSPACE, Tangram, MERFISH) that reconstruct the anatomical dimension compressed out by bulk RNA-seq. A clinically optimal strategy may combine these — an IHC+RNA-seq hybrid in which PU.1 IHC quantifies total TAM density while RNA-seq provides the TME compositional axis is a parsimonious translation of the Maas framework to routine pathology workflows. The present study defines the boundary conditions that any of these future approaches must exceed.

## Data Availability

Analysis code will be deposited on GitHub upon publication.

