## Supplementary for "Single-Cell-Validated Transcriptomic Proxies for the Maas Meningioma Microenvironment Risk Continuum: An NF2-Dependent Signal Attenuated Below Detectability in Bulk RNA-seq"

### Supplementary Materials

#### Supplementary Table S1. Microglia and Macrophage Gene Sets

**Microglia gene set (15 genes)**: TMEM119, P2RY12, P2RY13, GPR34, SLC2A5 (Maas core markers); CX3CR1, HEXB, SALL1, CSF1R, TREM2, OLFML3, SELPLG, SPI1, SIGLEC8, AIF1 (expanded microglial identity genes).

Note: SIGLEC8 was the available SIGLEC family member in the harmonized expression matrix; the originally intended SIGLECH (microglial-specific siglec) was not present. TREM2 has bidirectional roles in homeostatic and disease-associated microglia (DAM); it was included based on its canonical homeostatic microglial identity status [10] and higher expression in microglia-like TAMs in the Maas snRNA-seq data. SPI1 (PU.1) is a pan-myeloid transcription factor expressed by both microglia and macrophages; its placement in the microglia set reflects the Maas finding that PU.1 percentage is highest in microglia-rich, low-risk tumors [8] but partially inflates the microglia score with total myeloid content. HP (haptoglobin) was included per Maas et al. [8] as an acute-phase marker of activated macrophages rather than a lineage-defining gene. CD83 is an activation marker shared by dendritic cells and activated macrophages; in the Maas snRNA-seq data it marked a subset of MC-mal-enriched macrophages.

**Macrophage gene set (17 genes)**: C3, F10, EMILIN2, F5, GDA (Maas core markers); HP, SPP1, CD163, MARCO, FOLR2, MSR1, CD68, ITGAM, CCL2, TGFB1, IL10, CD83 (expanded tumor-supportive/infiltrating macrophage markers).

**Excluded from primary analysis**: MKI67, TOP2A (proliferation markers — Ki-67 signal in meningioma is contributed by both tumor and immune cells [14]), SELL (lymphocyte marker).

#### Supplementary Table S2. Cohort Characteristics

| Characteristic | Full cohort (N=968) | Survival cohort (N=101) |
| --- | --- | --- |
| **Dataset** |  |  |
| Baylor (GSE136661) | 160 (16.5%) | 17 (16.8%) |
| HKU/UCSF | 302 (31.2%) | 0 (0%) |
| UCSF 2018 | 42 (4.3%) | 0 (0%) |
| UCSF 2022 | 185 (19.1%) | 15 (14.9%) |
| UW/FHCC | 279 (28.8%) | 69 (68.3%) |
| **WHO grade** |  |  |
| Grade I | 121 (12.5%) | 34 (33.7%) |
| Grade II | 32 (3.3%) | 52 (51.5%) |
| Grade III | 7 (0.7%) | 15 (14.9%) |
| Not available | 808 (83.5%) | 0 (0%) |
| **Recurrence events** | — | 73 (72.3%) |
| **Median follow-up (months)** | — | 110.2 |

#### Supplementary Table S3. Pseudo-Bulk Validation: TAM Classification by MC Class

| MC class | N TAMs | Microglia-like | Macrophage-like | Microglia % |
| --- | --- | --- | --- | --- |
| ben-1 | 8,520 | 4,064 | 4,456 | 47.7% |
| ben-2 | 663 | 242 | 421 | 36.5% |
| int-A | 4,605 | 1,511 | 3,094 | 32.8% |
| int-B | 612 | 218 | 394 | 35.6% |
| mal | 1,379 | 199 | 1,180 | 14.4% |

#### Supplementary Table S4. Schoenfeld Residual Tests for Proportional Hazards Assumption

| Model | Variable | Chi-squared | p |
| --- | --- | --- | --- |
| Univariable | Ratio (z-scored) | 1.17 | 0.28 |
|  | Global | 1.17 | 0.28 |
| Multivariable | Ratio (z-scored) | 0.83 | 0.36 |
|  | WHO grade | 0.07 | 0.79 |
|  | Global | 0.98 | 0.61 |

#### Supplementary Table S5. Nassiri MG Proxy Assignment Distribution

| MG proxy group | N | % | Mean ratio | SD |
| --- | --- | --- | --- | --- |
| MG3-Hypermetabolic | 670 | 69.2 | +0.17 | 0.95 |
| MG4-Proliferative | 200 | 20.7 | -0.25 | 1.02 |
| MG2-NF2wt/Benign | 97 | 10.0 | -0.62 | 0.92 |
| MG1-Immunogenic | 1 | 0.1 | -1.03 | — |

Note: The expected distribution based on Nassiri et al. [6] is approximately 25% per group. The 69.2% assignment to MG3-Hypermetabolic indicates that Hallmark pathway-based proxy scoring does not reconstruct the Nassiri molecular groups. These results should not be interpreted as meaningful concordance data; the original gene signatures are required for valid molecular group assignment.

#### Supplementary Table S6. Extended Sensitivity Analyses (R3)

| Analysis | Model | HR | 95% CI | p | C-index |
| --- | --- | --- | --- | --- | --- |
| **Age adjustment** |  |  |  |  |  |
| WHO + age | age | 1.02 | 1.00-1.03 | 0.049 | 0.612 |
| WHO + age + ratio | ratio | 0.99 | 0.78-1.25 | 0.91 | 0.615 |
| **Institution stratification** |  |  |  |  |  |
| Stratified by site (UV) | ratio | 0.92 | 0.72-1.17 | 0.49 | — |
| Stratified by site (MV+WHO) | ratio | 1.03 | (CI not reported - see step02_survival.R) | 0.81 | — |
| UW/FHCC only (N=69) | ratio | 0.92 | 0.71-1.19 | 0.54 | — |
| **NF2 expression proxy** |  |  |  |  |  |
| NF2-low (N=51, 36 ev) UV | ratio | 0.68 | 0.46-1.01 | 0.056 | — |
| NF2-low MV+WHO | ratio | 0.74 | 0.48-1.15 | 0.19 | — |
| NF2-high (N=50, 37 ev) UV | ratio | 0.98 | 0.71-1.34 | 0.89 | — |
| **Composition-corrected** |  |  |  |  |  |
| Residualized ratio (UV) | ratio_resid | 0.94 | 0.75-1.18 | 0.59 | — |
| Residualized ratio (MV+WHO) | ratio_resid | 0.98 | 0.77-1.23 | 0.84 | — |
| **Component scores** |  |  |  |  |  |
| Microglia score (UV) | microglia | 0.91 | 0.73-1.14 | 0.42 | 0.538 |
| Macrophage score (UV) | macrophage | 0.97 | 0.77-1.22 | 0.77 | 0.500 |
| **Chen 34-gene benchmark** |  |  |  |  |  |
| Chen panel (UV, 32/34 genes) | chen_34 | 0.92 | 0.72-1.17 | 0.49 | 0.552 |
| Chen panel (MV+WHO) | chen_34 | 0.83 | 0.65-1.06 | 0.13 | — |
| **C-index formal comparison** |  |  |  |  |  |
| WHO vs WHO+ratio | delta-C | +0.022 | — | 0.20 | — |
| WHO vs WHO+macro | delta-C | -0.018 | — | 0.32 | — |

Note: NF2 expression proxy uses median split of NF2 mRNA expression; NF2-low is enriched for but does not confirm NF2-mutant tumors. Composition-corrected ratio = residuals after regressing ratio on EPIC macrophage score. Chen 34-gene panel from Chen et al. [4], validated at AUC 0.81 (n=1,856); 32/34 genes available (CCN1 and LINC02593 absent from expression matrix). All HRs per 1 SD increase. NF2-low EPV: 36 events / (ratio + WHO) = 18 per covariate.

#### Supplementary Methods: Attenuation Analysis Derivation

The expected attenuation of the Maas IHC reference HR (2.00, from PU.1/TAM density in n=1,378, 321 events [8]) to the observable bulk RNA-seq HR was computed using two compounding mechanisms.

**1. Classical errors-in-variables attenuation.** Under the measurement-error (regression-dilution) model, log hazard ratios attenuate linearly with reliability:

$$log(\text{HR}_{\text{obs}})=\lambda\cdot log(\text{HR}_{\text{true}})$$

where lambda is the reliability of the proxy, operationalized as the coefficient of determination (r^2^) of the ssGSEA ratio against the Maas risk construct. Two reliability estimates bound the plausible range:

- **Conservative lower bound**: r^2^ = 0.22, from the Spearman correlation between the pseudo-bulk ssGSEA ratio and the ordinal Maas MC risk class (r = -0.47, n=25). This estimate captures the ratio’s capacity to resolve the coarse-grained 5-level ordinal risk class rather than the underlying continuous biology, and therefore understates reliability.
- **Strict upper bound**: r^2^ = 0.49, from the strict pseudo-bulk correlation with single-cell microglia proportion (r = 0.70, n=25, circularity-controlled). This estimate targets the continuous biology directly.

Under r^2^ = 0.22, expected HR = 2.00^0.22 approximately 1.17 on the multiplicative scale; equivalently, log(HR_obs) = 0.22 x log(2.00) = 0.153, giving HR_obs approximately 1.17. Under r^2^ = 0.49, HR_obs approximately 1.40. A commonly-applied alternative specification uses the square root of reliability as the attenuation factor (lambda = sqrt(r^2^)); under that parameterization, the chain would yield HR approximately 1.39 (sqrt(0.22)) to HR approximately 1.63 (sqrt(0.49)). The main text reports the range HR approximately 1.24-1.40 as a conservative midpoint across both reliability estimates and both parameterizations, with the lower values combining conservative reliability with NF2 dilution.

**2. NF2-wildtype dilution.** Because the Maas risk continuum is defined within NF2-mutant meningiomas [8], the observable effect in an unselected cohort is a weighted average of the NF2-mutant effect (HR_true) and the NF2-wildtype effect (assumed HR = 1, no effect). On the log scale:

$$log(\text{HR}_{\text{pooled}})=\pi_{\text{NF2-mut}}\cdot log(\text{HR}_{\text{true}})$$

where pi is the NF2-mutant fraction. Grade-weighted NF2-wildtype prevalence was derived from published NF2 mutation rates by WHO grade (~50% NF2-mutant in WHO 1, ~75% in WHO 2, ~80% in WHO 3; Choudhury 2022 [3]; Maas 2026 [8]) applied to the MENENV cohort grade distribution (WHO 1: 34, WHO 2: 52, WHO 3: 15 of n=101). The resulting NF2-wildtype fraction is approximately 30-45% across plausible grade-specific rates, bracketing the 35% midpoint used in the main text.

Combining both mechanisms under the conservative parameterization (r^2^ = 0.22, NF2-wildtype = 45%, sqrt(r^2^) attenuation): HR_obs approximately (2.00^sqrt(0.22))^0.55 approximately 1.20. Under the strict parameterization (r^2^ = 0.49, NF2-wildtype = 30%, linear attenuation): HR_obs approximately 2.00^(0.49 x 0.70) approximately 1.40. The reported range of HR 1.24-1.40 spans these boundary conditions.

**3. Power.** With 73 events and alpha=0.05 two-sided, power to detect a continuous predictor was computed using the Schoenfeld (1983) formula under the observed ratio variance (SD = 1 after z-scoring). Power is approximately 15% at HR 1.24, approximately 29% at HR 1.39, approximately 40% at HR 1.50. At 80% power, approximately 480 events would be required to detect HR 1.24 (the grade-weighted, NF2-diluted expected effect). Code for the attenuation range and power computation is included in the analysis repository.
